# Comparing Physicians’ Assessments of Context-specific AI-powered clinical reasoning assistant with General-Purpose AI agent: A Prospective Multi-Site Physician Evaluation of VITA versus ChatGPT in India and Bangladesh

**DOI:** 10.64898/2026.04.30.26351194

**Authors:** Charuta Mandke, Hemendra Kumar Agrawal, Bhartendu Bharti, Mayank Chansoria, Gourav Gupta, Sumit Kumar Rawat, Nirmal Kanti Sarkar, Anil Singh, P.S. Suji, Shweta Walia, and the VALID Consortium

## Abstract

**Background:** Healthcare providers in low- and middle-income countries (LMICs) are increasingly relying on Artificial Intelligence (AI) tools, yet most available AI assistants are general-purpose systems not designed for the specific clinical, epidemiological, and resource contexts of these settings. There is no evidence, from physicians’ assessments, on whether clinical reasoning support from purpose-built, context-specific and retrieval-augmented AI tools can outperform general-purpose AI agents.

**Methods:** We conducted a prospective multi-site validation study enrolling 37 physicians across India and Bangladesh. Each physician evaluated two AI tools (a) VITA (Validated Intelligence for Treatment and Assessment), a purpose-built (context-specific and retrieval-augmented) clinical reasoning AI assistant trained on India-specific guidelines, antimicrobial resistance patterns, and formulary constraints, and (b) ChatGPT Plus (version 5.2), a leading general-purpose AI assistant on six hypothetical clinical case vignettes (three predefined, three physician-selected). Evaluations were scored across six dimensions (differential diagnosis, clinical workup, treatment recommendation, dosing, clinical decision-making, and evidence quality) on a 1–5 Likert scale, yielding 444 observations. Analyses included paired t-tests, Wilcoxon signed-rank tests, and multivariate regressions with robust standard errors.

**Results:** VITA scored significantly higher than ChatGPT across all six evaluation dimensions. The mean composite score (sum of all dimensions, maximum = 30) was 25.4 for VITA versus 22.3 for ChatGPT (difference = +3.1 points, t = 8.31, p < 0.001). The largest advantage was in evidence quality (VITA: 4.46 vs. ChatGPT: 3.14, a 42% relative gap). VITA’s advantage was consistent across both predefined and doctor-defined hypothetical cases and was robust to controls for physician demographics, case type, and evaluation order in multivariate regression (coefficient = +3.08, p < 0.001).

**Conclusions:** In this first systematic head-to-head physician evaluation of a purpose-built clinical reasoning AI assistant versus general-purpose AI in an LMIC setting, physicians consistently rated the context-specific tool as superior. These findings suggest that contextual relevance—including local guidelines, formulary constraints, and resistance patterns—matters for clinical AI adoption and quality in resource-limited settings.

## 1. Introduction

The ongoing global proliferation of artificial intelligence (AI) tools in healthcare represents one of the most consequential technological shifts in modern medicine. AI applications ranging from diagnostic imaging and medical scribes, to natural language-based clinical assistants are being developed, tested, and deployed at unprecedented scale across both high-income and low- and middle-income country (LMIC) healthcare systems (Ciecierski-Holmes et al. 2022; Alami et al. 2020). In settings where access to care and clinical capacity remain severely constrained, digital health technologies and AI have the potential to be transformative. (WHO 2020).

The promise of AI in healthcare must be weighed against long-standing and well-documented challenges in the quality of clinical care in resource-limited settings. An estimated 5.7 to 8.4 million deaths annually in LMICs are attributable to poor-quality care, and poor quality imposes productivity losses of $1.4 to $1.6 trillion per year (National Academies of Sciences 2018). Studies across countries from India to Tanzania have documented wide variation in provider competence, with the “know–do gap”, defined as the gap between providers’ knowledge when assessed using standardized vignettes and what they actually do in clinical practice, emerging as a persistent and pervasive challenge (Das and Hammer 2014; Das et al. 2012; Mohanan et al. 2015). This gap reflects individual skill deficits such as the inability to apply knowledge in clinical settings as well as systemic problems including inadequate supervision, low motivation, lack of incentives, poor infrastructure, supply chain failures for drugs and diagnostics, and insufficient accountability mechanisms (Kruk et al. 2018; Leslie et al. 2017).

Against this backdrop, many healthcare providers in developing countries have begun using ChatGPT and other commercially available large language models (LLMs) to assist with diagnosis, treatment planning, and clinical decision-making (Si et al. 2024; Khosla 2023). A cross-national study found that interest in ChatGPT was significantly higher in countries with lower physician density, suggesting that AI may be filling perceived gaps in clinical support where human expertise is scarce (Eid et al. 2025). Reports in both the medical literature and mainstream media have documented the rapid, often informal adoption of general-purpose AI chatbots by clinicians across South Asia, sub-Saharan Africa, and Latin America (Agrawal et al. 2024). This organic uptake is especially notable because it has occurred largely outside formal health system channels, without structured training on AI capabilities and limitations, and without guardrails on whether these clinicians can evaluate or critically appraise AI-generated clinical recommendations.

While the enthusiasm for AI-assisted clinical care is understandable, the use of general-purpose AI tools in LMIC clinical settings raises significant concerns. General-purpose LLMs are not trained on local clinical guidelines, do not account for regional drug availability or formulary constraints, are often unaware of local antimicrobial resistance patterns, and may generate recommendations that are clinically sound in a high-income context but inappropriate or infeasible in resource-limited settings (Si et al. 2024; Agrawal et al. 2024). Contextually inappropriate recommendations are a threat because general-purpose LLMs do not account for epidemiological profiles of LMIC populations, including the burden of tropical infections, co-morbidity patterns, and the prevalence of conditions rarely seen in the training data of most LLMs.

There is therefore a pressing need for AI-powered clinical reasoning tools to that are specifically designed for LMIC healthcare contexts: tools that are contextually relevant, account for resource constraints and the availability of diagnostics and drugs, incorporate local epidemiological data and antimicrobial resistance patterns, and are aligned with national clinical guidelines. Equally important is the need for rigorous, physician-centered evaluations that compare such purpose-built (context-specific and retrieval-augmented) clinical reasoning AI assistant tools against the general-purpose AI tools that clinicians are already using informally.

## 2. Literature on Digital Clinical Decision Support, AI, and LLMs in Low-Resource Settings

Digital tools for clinical decision support have long been attractive in low- and middle-income countries (LMICs) because they offer a possible way to standardize care where clinical training, supervision, diagnostics, and specialist access are uneven. The evidence base for health worker decision support through digital health intervention is, however, stronger for process outcomes than for downstream health outcomes (World Health Organization 2019).

Most available evidence in the literature is still centered on structured, guideline-based digital clinical decision support algorithms (CDSAs) rather than on fully generative AI. Such CDSAs in low-resource settings typically combine digital algorithms and point-of-care diagnostics to improve evidence-based treatment decisions by incorporating symptoms, signs, test results, and contextual information such as seasonality and local disease incidence (Tamrat et al. 2020; World Health Organization 2020). While CDSAs can improve process quality or adherence to guidelines, they do not on their own resolve implementation and adoption related issues such as staffing, motivation, supervision, or workflow constraints (Tallarico et al. 2025).

The rapid expansion of AI in healthcare has shifted attention away from guideline-based CDSAs toward the broader potential of large language models (LLMs) to support clinical decision-making across diverse settings. LLMs are particularly salient because they may address some of the limitations inherent in earlier rule-based systems. Unlike static decision trees, they can respond to free-text queries, accommodate incomplete or evolving case descriptions, and provide multilingual, context-sensitive guidance. These capabilities are especially relevant in low-resource settings, where clinicians frequently encounter heterogeneous presentations, limited diagnostic capacity, and clinical decisions that do not map neatly onto rigid algorithms. The flexibility of LLMs, in principle, aligns well with these realities.

At the same time, the emerging empirical literature offers a mixed picture. In a randomized clinical trial among U.S. physicians in general medical specialties, access to an LLM did not lead to significant improvements in diagnostic reasoning relative to conventional resources (Goh et al. 2024). In contrast, a subsequent randomized controlled trial, which focused on open-ended management reasoning tasks derived from expert-developed clinical vignettes, found that GPT-4 assistance improved physician performance on complex decision-making tasks in a simulated setting (Goh et al. 2025). Taken together, these findings suggest that the value of LLMs may depend critically on the type of clinical task and context in which they are deployed.

For LMICs specifically, direct evidence on LLM-assisted clinical reasoning is only now emerging. A 2026 study based in Rwanda developed a dataset of 5,609 real clinical questions contributed by 101 community health workers across four districts and found that several general-purpose LLMs outperformed local clinicians on expert-rated quality metrics; the study also reported that model performance remained strong when prompted in *Kinyarwanda* (Ntirenganya et al. 2026). A second 2026 randomized controlled trial in Pakistan found that physicians who received AI-literacy training and then used LLM assistance achieved substantially higher diagnostic reasoning scores than physicians using conventional resources alone, with no time penalty (Qazi et al. 2026). While the evidence from these settings is suggestive, the process outcomes in these studies reflect the current state of adoption and implementation of AI tools: the Rwanda study evaluated question–answer exchanges rather than patient outcomes, and the Pakistan trial was vignette-based rather than embedded in routine clinical care.

The implementation science of clinical LLMs remains nascent, particularly in LMIC settings, where evidence on their effects on patient outcomes is still emerging. In this context, a central question for the adoption of AI in healthcare is how clinicians themselves evaluate different forms of AI assistance when engaged in real clinical reasoning. Do they view these systems as useful, trustworthy, contextually appropriate, and safe? Do they integrate smoothly into existing workflows and resource constraints?

Most existing studies in LMICs have taken a narrower approach—comparing a single digital tool to usual care, benchmarking model outputs against expert judgment, or focusing on performance in specific tasks. Far less attention has been paid to how physicians compare competing AI systems directly.

This gap is consequential. Studies that elicit physicians’ comparative assessments of alternative AI tools can offer a distinct and policy-relevant contribution. They speak directly to questions of adoption, usability, and contextual fit—factors that are likely to be as important as benchmark accuracy in determining whether AI meaningfully improves clinical decision-making in low-resource settings (Ciecierski-Holmes et al. 2022; Ashkenazi et al. 2025).

## 3. Study Design and Methods

### 3.1 Study Objective

This study was designed to address a specific question: how do physicians assess the relative performance of an AI-powered clinical decision support tool purpose-built for India and similar LMIC healthcare settings, compared to a leading general-purpose AI assistant? The two tools evaluated were VITA (Validated Intelligence for Treatment and Assessment) and ChatGPT Plus (version 5.2).

### 3.2 The VITA System

VITA is a clinical reasoning AI assistant that uses a retrieval-augmented generation (RAG) architecture to generate responses grounded in curated, locally relevant evidence. Unlike general-purpose large language models that generate responses from parametric knowledge alone, VITA’s outputs are anchored to a structured knowledge corpus assembled specifically for the Indian clinical context. VITA’s structured knowledge base is created with a proprietary curation process that includes quality assessment and metadata tagging that is frequently updated. The knowledge base used in VITA’s RAG architecture incorporates Indian national clinical guidelines (including national guidelines, national antibiotic stewardship programs, and specialty society recommendations), India’s national antimicrobial resistance surveillance data (NARS-Net), and locally relevant clinical literature.

For each clinical query, VITA executes a multi-phase pipeline for retrieval and generation process. Each submitted clinical query undergoes clinical entity extraction and semantic enrichment before retrieval. Relevant documents are then identified from the knowledge corpus using semantic similarity; retrieved evidence is ranked and synthesized with explicit source attribution. The final response integrates retrieved evidence with India-specific drug formulary data and antimicrobial resistance profiles.

### 3.3 Study Population and Setting

This prospective validation study enrolled 37 physicians across India and Bangladesh. Participating physicians represented multiple specialties, including ophthalmology, internal medicine, anesthesiology, otolaryngology and critical care, and were located in nine cities: Agra, Deoghar, Dhaka, Indore, Jabalpur, Mumbai, Raipur, Rajkot, and Sagar. The study was administered online using a structured Qualtrics survey tool.

### 3.4 Evaluation Framework

This study relies on a within-physician paired design in which each respondent serves as their own control. The evaluation was conducted by physicians and experts recruited as a convenience sample to include expertise in a broad range of clinical specialties. Each physician evaluated both AI tools on six hypothetical clinical case vignettes: three predefined cases and three doctor-defined cases (written by the evaluating physician based on cases from their own clinical practice). For each case, the physician queried both VITA and ChatGPT with the same clinical scenario and scored each tool’s response across six dimensions on a 1–5 Likert scale: (1) differential diagnosis, (2) clinical workup, (3) treatment recommendation, (4) dosing recommendation, (5) clinical decision-making quality, and (6) quality of evidence cited. Each physician respondent relied on their own expert knowledge to benchmark performance of both tools for assessment on the 5-point Likert scale. Blinding of participants to whether they were using GPT or VITA was not possible; participants logged into each of the tools using subscriber login credentials provided to them. A composite total score was computed as the sum across all rated dimensions (maximum = 30). This yielded 444 paired physician–tool–case observations (37 physicians × 6 cases × 2 tools). No patients or human subjects were involved in this study; none of the cases described by physician respondents included any patient or identifying information.

### 3.5 Statistical Analysis

Descriptive statistics were computed for physician characteristics. Differences between VITA and ChatGPT were assessed using paired t-tests for mean scores at the case-by-dimension level and for composite scores, as well as Wilcoxon signed-rank tests for paired differences. To assess whether the observed VITA advantage could be explained by physician characteristics, case type, or evaluation order, we estimated OLS and Fixed effects regressions of the composite total score on a VITA indicator, controls for case type (predefined vs. doctor-defined), case order (1–6), physician gender, and physician age group, with heteroskedasticity-robust (HC1) standard errors. As a robustness check, we also ran alternate regressions with physician-clustered standard errors; the results were identical to those reported in the main analysis. All analyses were conducted in Stata 16+.

To minimize conflicts of interest, all analyses were replicated using Claude Pro and ChatGPT Pro, where both AI agents were provided with anonymized raw data and asked to produce tables and regression results, and then verified against each other.

## 4. Results

### 4.1 Physician Characteristics

Table 1 presents descriptive statistics on the 37 participating physicians. The sample included physicians from nine cities across India and one site in Bangladesh (Dhaka), spanning multiple specialties and career stages.

**Table 1.** Descriptive Statistics of Participating Physicians.

| Category | Detail | N (%) |
| --- | --- | --- |
| Total Physicians |  | 37 |
| Total Evaluations | (37physicianX 6casesX 2tools) | 444 |
| Gender |  |  |
|  | Male | 23 (62.2%) |
|  | Female | 14 (37.8%) |
| Age Group |  |  |
|  | 25-30 | 17 (45.9%) |
|  | 30-40 | 11 (29.7%) |
|  | 40-50 | 8 (21.6%) |
|  | 50-60 | 1 (2.7%) |
| Medical Specialty |  |  |
|  | Otorhinolaryngology | 7 (18.9%) |
|  | Ophthalmology | 6 (16.2%) |
|  | Pulmonology | 6 (16.2%) |
|  | Anesthesiology | 5 (13.5%) |
|  | Internal Medicine | 5 (13.5%) |
|  | Oncology | 4 (10.8%) |
|  | Emergency Medicine | 2 (5.4%) |
|  | Microbiology | 2 (5.4%) |
| Practice Location |  |  |
|  | Agra | 5 (13.5%) |
|  | Deoghar | 7 (18.9%) |
|  | Dhaka | 6 (16.2%) |
|  | Indore | 6 (16.2%) |
|  | Jabalpur | 4 (10.8%) |
|  | Mumbai | 3 (8.1%) |
|  | Raipur | 4 (10.8%) |
|  | Rajkot | 1 (2.7%) |
|  | Sagar | 1 (2.7%) |

Most participating physicians (62.2%) were male and in the 25–40 age range (45.9%). Qualifications ranged from consultants and specialists to postgraduate residents and fellows. Respondents represented a mixed set of specialties, including Otorhinolaryngology (18.9%), followed by Pulmonology (16.2%), Ophthalmology (16.2%), Internal medicine (13.5%), Anesthesiology (13.5%), Oncology (10.8%), Emergency Medicine(5.4%) and Microbiology (5.4%).

### 4.2 Case Vignettes

Table 2 summarizes the structure of the hypothetical clinical case vignettes used in the study. Three predefined cases scenarios were standardized across all participants to ensure comparability, while three additional hypothetical clinical case scenarios were provided independently by each physician based on their own clinical practice, ensuring contextual validity and relevance to real-world clinical decision-making.

**Table 2.** Hypothetical Clinical Case Vignettes.

| Case Order | Case Type | Description |
| --- | --- | --- |
| Case 1 | Predefined | Standardized clinical scenario provided to all physicians:<br>75year male with heart failure and reduced ejection fraction. How can Entresto (sacubitril / valsartan) be used? |
| Case 2 | Doctor-Defined | Examples of clinical scenario provided by evaluating physicians. The questions are included verbatim without editing for clarity:<br>30 year old female diagnosed with Carcinoma Cervix Ib3 in 2nd trimester of pregnancy. what should be the line of management? [sic]<br>27 years-old male came with recurrent chest infection, cough with productive sputum production. On examination, clubbing is present in all fingers & toes. HbA1C 9.5%. How to approach to this patient for further evaluation to reach diagnosis & treatment? [sic] |
| Case 3 | Predefined | Standardized clinical scenario provided to all physicians:<br>68 year old diabetic, hospital day 5 post-abdominal surgery, developing fever, hypotension (BP 88/52), lactate 3.2. Currently on piperacillin-tazobactam. Culture pending (48 hours). ICU has limited resources - no procalcitonin, basic ventilator only. Recommend treatment protocol for this patient.: |
| Case 4 | Doctor-Defined | Examples of clinical scenario provided by evaluating physicians. The questions are included verbatim without editing for clarity:<br>How to assess treatment response to steroid in a case of ABPA [sic]<br>A 65 years old female patient complaining of ringing sensation in both ears. How to proceed? [sic] |
| Case 5 | Predefined | Standardized clinical scenario provided to all physicians:<br>35y lady with UTI in Noida. Which antibiotic? |
| Case 6 | Doctor-Defined | Examples of clinical scenario provided by evaluating physicians. The questions are included verbatim without editing for clarity:<br>A 35 year old male presented in eye OPD with diminution of vision in LE -Hand movement PR accurate, with yellowish reflex in LE since 10 days with conjunctival congestion. episodes of similar conjunctival congestion 1 year back. what can be differential diagnosis of this patient [sic]<br>Make a plan for evaluation and management of a previously healthy 42-year-old lady presenting with massive haemoptysis for 2 hours. [sic] |

*Note: Each physician evaluated both VITA and ChatGPT on each hypothetical case, querying both tools with the identical clinical scenario.*

### 4.3 Dimension-Level Comparisons

Figure 1 presents the mean physician scores for VITA and ChatGPT across all six evaluation dimensions, pooled across all cases. VITA scored higher than ChatGPT on every dimension. The largest absolute difference was observed in the quality of evidence cited (VITA: 4.5 vs. ChatGPT: 2.91, a difference of 1.6 points on the 1–5 scale, representing a 54% relative gap). Substantial differences were also observed for clinical decision-making (VITA: 4.37 vs. ChatGPT: 3.89), dosing recommendations (VITA: 4.27 vs. ChatGPT: 3.77), treatment recommendations (VITA: 4.29 vs. ChatGPT: 3.82), clinical workup (VITA: 4.27 vs. ChatGPT: 3.84), and differential diagnosis (VITA: 4.36 vs. ChatGPT: 3.98).

**Figure 1.**
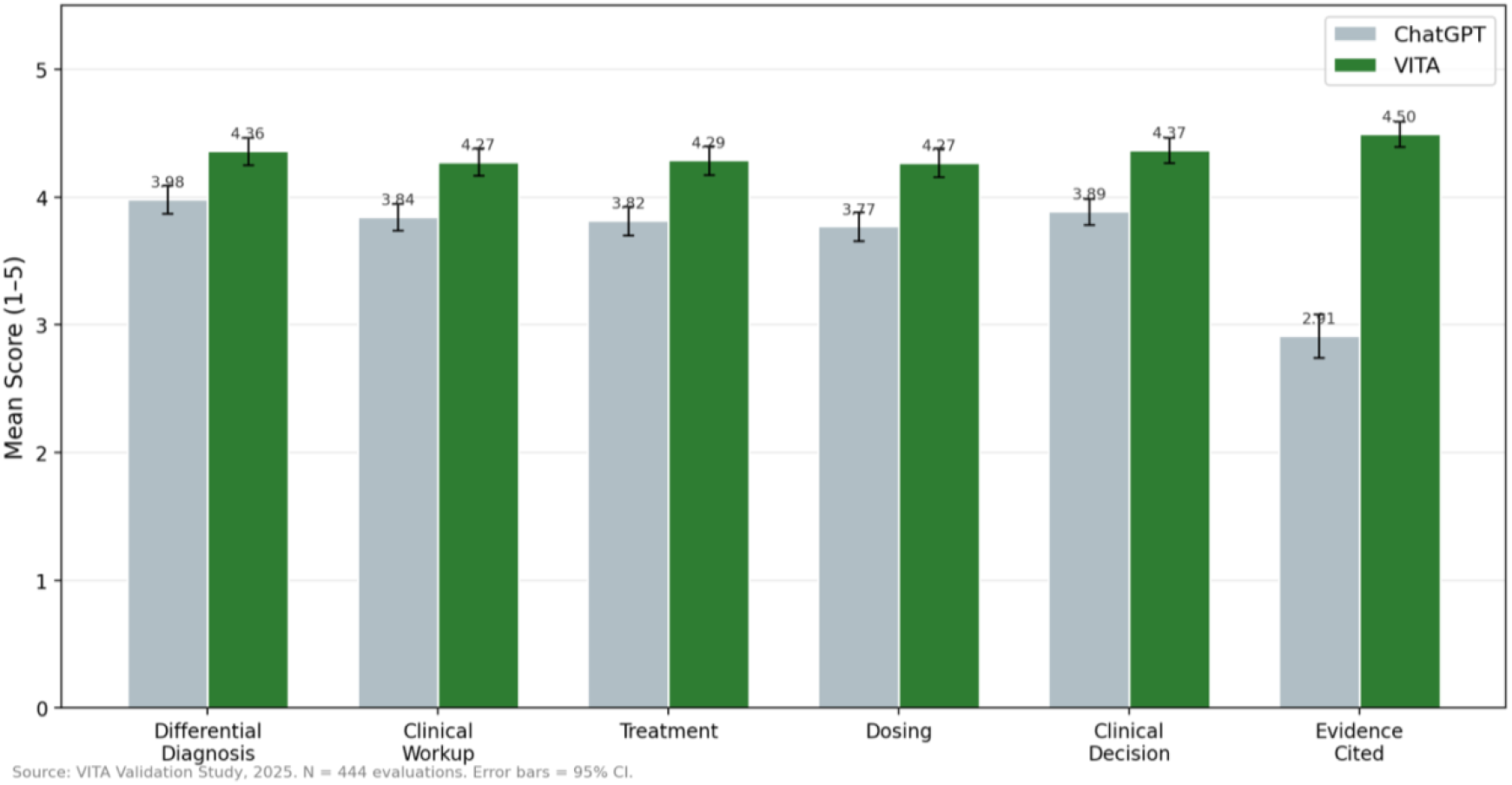
VITA vs. ChatGPT: Mean Scores Across All Dimensions.

The pattern of results is notable: while VITA’s advantage was present across all dimensions, it was most pronounced in evidence quality, the dimension most directly connected to the ability to cite relevant, locally appropriate clinical guidelines and research. This is consistent with VITA’s design, which incorporates India-specific evidence sources. The smaller gap in differential diagnosis (0.38 points) suggests that general-purpose LLMs can generate plausible differential diagnoses, but struggle more with context-specific treatment, dosing, and evidentiary support.

### 4.4 Composite Score Comparison

Figure 2 presents the composite assessment score, computed as the sum of physician ratings across all six dimensions (differential diagnosis, clinical workup, treatment, dosing, clinical decision-making, and evidence cited), for VITA and ChatGPT. The distributional composition in Panel (a) displays overlapping histograms with kernel density curves, showing VITA distribution visibly shifted to the right (higher scores) relative to ChatGPT. Panel (b) shows the mean composite scores with 95% confidence intervals. VITA received a significantly higher mean composite score than ChatGPT (26.1 ± 3.9 vs. 22.2 ± 4.0; difference = 3.9 points; t = 10.36, p < 0.001). A Wilcoxon signed-rank test on within-physician paired differences (W = 2,601, p < 0.001) that accounts for the paired structure of the data, because each physician rated both tools on the same case indicates that physicians consistently rated VITA’s clinical recommendations higher than ChatGPT’s across the full range of evaluated dimensions.

**Figure 2.**
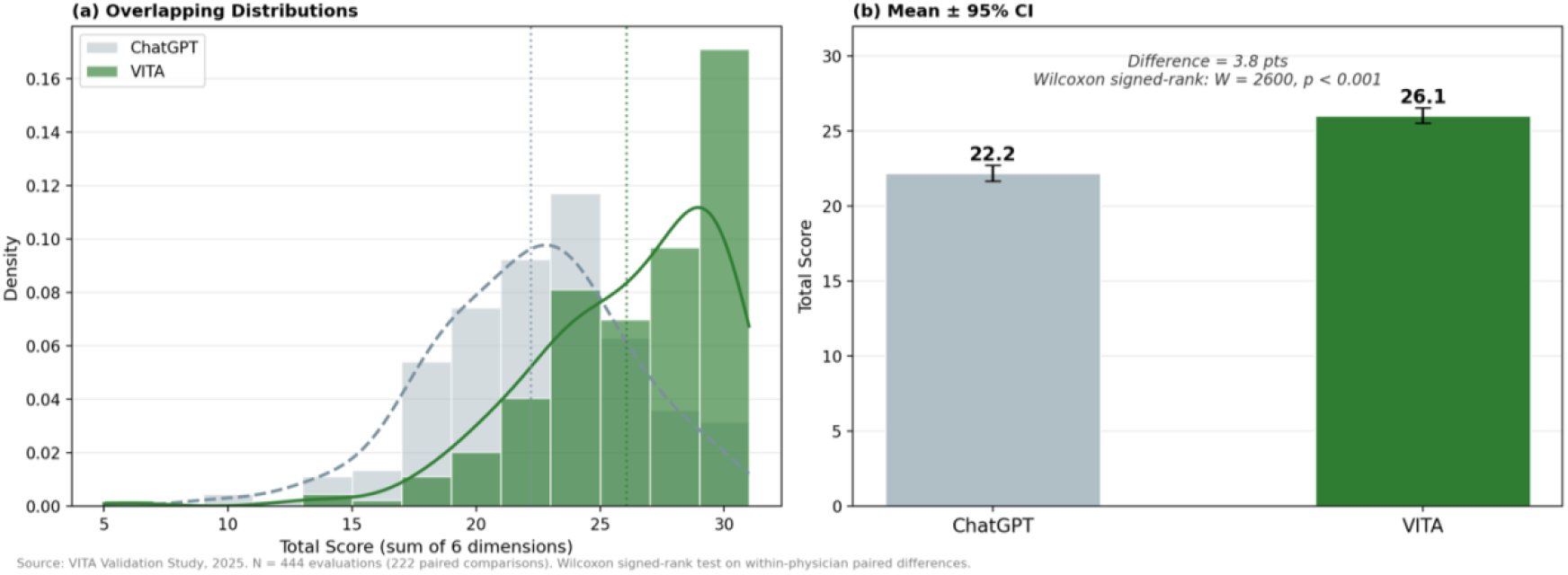
Composite Assessment Index: VITA vs. ChatGPT.

### 4.5 Results by Case Type

An important question is whether VITA’s advantage holds across both predefined (standardized) and doctor-defined (physician-selected) cases. Figure 3 presents dimension-level mean scores separately for each case type. VITA outperformed ChatGPT on every dimension of physician assessments for both predefined and doctor-defined cases. For predefined cases, the VITA advantage was evident in differential diagnosis (4.35 vs. 3.93), clinical workup (4.27 vs. 3.81), treatment (4.32 vs. 3.79), dosing (4.43 vs. 3.88), clinical decision-making (4.34 vs. 3.86), and evidence cited (4.56 vs. 2.92), with all differences statistically significant (p < 0.001 for all dimensions). For doctor-defined cases, where physicians selected clinical scenarios from their own practice, VITA’s advantage relative to ChatGPT was similarly consistent on differential diagnosis (4.37 vs. 4.04, p = 0.003), clinical workup (4.28 vs. 3.87, p < 0.001), treatment (4.25 vs. 3.84, p < 0.001), dosing (4.11 vs. 3.66, p < 0.001), clinical decision-making (4.40 vs. 3.91, p < 0.001), and evidence cited (4.43 vs. 2.91, p < 0.001).

**Figure 3.**
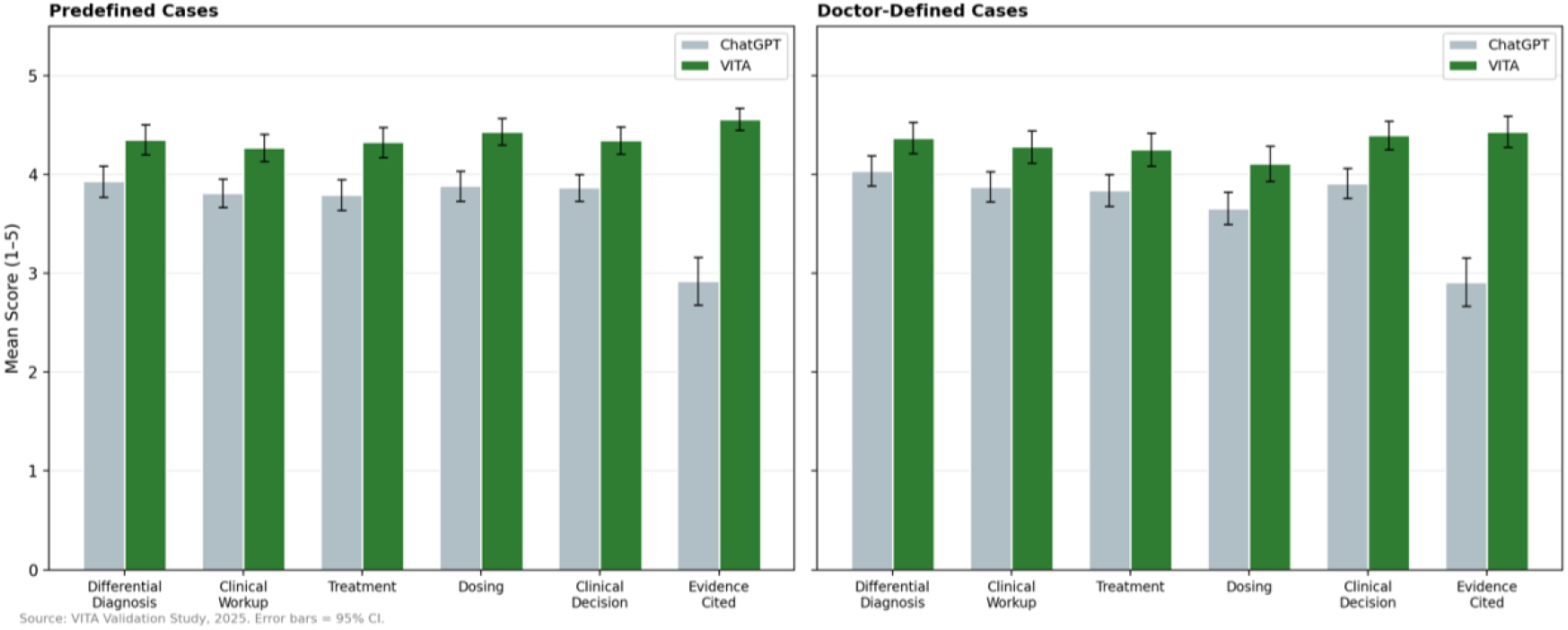
VITA vs. ChatGPT by Case Type (Predefined vs. Doctor-Defined.

The largest gap in both case types was on the Evidence Cited dimension, where VITA scored approximately 1.5 points higher than ChatGPT. The consistency of VITA’s advantage across both standardized and physician-selected cases strengthens the conclusion that the results are not an artifact of a particular case design. That VITA performs well on doctor-defined cases - which are, by definition, more heterogeneous and less predictable than standardized vignettes - suggests that the tool’s contextual knowledge generalizes to the kinds of clinical scenarios physicians encounter in practice.

### 4.6 Multivariate Regression Analysis

To assess whether the observed VITA advantage could be explained by physician demographics, case characteristics, or evaluation order effects, we conducted regression analyses of the composite total score on a VITA indicator and a set of control variables including whether the case was predefined or doctor-defined, order of cases (to account for learning by the LLM or the doctor), as well as demographic characteristics.

Table 3 presents results from two regression specifications examining the determinants of the composite assessment score. In Model 1 (OLS with robust standard errors), the VITA indicator was the strongest predictor, with physicians rating VITA 3.85 points higher than ChatGPT on the composite score (β = 3.85, SE = 0.37, p < 0.001). Case order was positively associated with scores (β = 0.29, SE = 0.11, p = 0.008), suggesting that physicians gave modestly higher ratings to both tools as they progressed through the evaluation. Neither case type, physician gender, nor age group was a significant predictor of total scores, indicating that the VITA advantage was not driven by observable physician characteristics or by whether the case was predefined or doctor-defined.

**Table 3.** Regression analyses of physician assessment total scores.

|  | Model 1: OLS |  |  | Model 2: Fixed Effects |  |  |
| --- | --- | --- | --- | --- | --- | --- |
|  | Coeff<br>(Rob. SE) | t-statistic | p-value | Coeff<br>(Rob. SE) | t-statistic | p-value |
| VITA (=1; ChatGPT =0) | 3.847***<br>(0.368) | 10.45 | <0.001 | 3.847***<br>(0.315) | 13.12 | <0.001 |
| Predefined Case | 0.499<br>(0.397) | 1.26 | 0.209 | 0.499<br>(0.084) | 1.58 | 0.114 |
| Case Order (1-6) | 0.292***<br>(0.110) | 2.66 | 0.008 | 0.292***<br>(0.084) | 3.47 | <0.001 |
| Male Physician | 0.369<br>(0.395) | 0.93 | 0.351 |  |  |  |
| Age 30-40 (ref: 25-30) | -1.011**<br>(0.433) | -2.34 | 0.019 |  |  |  |
| Age 40-50 (ref: 25-30) | -0.236<br>(0.569) | -0.42 | 0.678 |  |  |  |
| Physician FE |  | N |  |  | Y |  |
| R2 |  | 0.2181 |  |  | 0.5412 |  |
| Adj R2 |  | 0.2074 |  |  | 0.4969 |  |
| N |  | 444 |  |  | 444 |  |

Notes: Dependent variable is *TotalScore\_i* (sum of physician scores across rated dimensions, max = 30). Model 1 shows OLS estimates with HC1 heteroskedasticity-robust standard errors in parentheses, Model 2 shows results from fixed effects regression. Reference categories: ChatGPT (tool), Doctor-defined (case type), Female (gender), Age 25–30. \*\*\* $p < 0.01$ , \*\* $p < 0.05$ , \* $p < 0.10$ .

We also present findings from a model that includes physician fixed effects with robust standard errors to absorb all time-invariant physician-level heterogeneity by including physician-specific intercepts, thereby eliminating potential confounding from unobserved physician traits such as clinical experience, risk tolerance, or rating tendencies. The VITA coefficient remained virtually unchanged at 3.85 (SE = 0.29, p < 0.001), and the case order effect was also robust (β = 0.29, SE = 0.08, p < 0.001). The R^2^ increased substantially to 0.54, reflecting the large share of variance attributable to physician-level differences in scoring. The stability of the VITA coefficient across the two specifications confirms that the observed advantage of VITA over ChatGPT is not an artifact of study design, physician composition or unobserved individual characteristics, but rather reflects a consistent within-physician preference for VITA’s clinical recommendations.

## 5. Discussion

This study provides the first systematic, multi-site, physician-evaluated head-to-head comparison of a purpose-built, context-specific and retrieval augmented, clinical reasoning AI assistant (VITA) against a leading general-purpose AI assistant (ChatGPT) in an LMIC setting. The results are clear and consistent: physicians across India and Bangladesh rated VITA significantly higher than ChatGPT on every dimension of clinical decision support quality, with the largest advantage emerging in the quality of evidence cited—the dimension most directly connected to safe, guideline-concordant care in resource-limited settings.

### 5.1 Contextual Relevance Matters

The central finding of this study, that a context-specific AI tool was rated higher than a general-purpose one in physician assessments, has important implications for the design and deployment of AI in LMIC healthcare. While general-purpose LLMs such as ChatGPT have demonstrated impressive capabilities in clinical reasoning tasks in high-income settings (Goh et al. 2024; Goh et al. 2025), our results suggest that these capabilities do not automatically translate into perceived clinical utility when physicians are evaluating recommendations for patients in resource-constrained environments. The gap is most pronounced in evidence quality, suggesting that clinicians seeking guidance prefer evidence that is aligned with local formularies, resistance data, and national guidelines.

### 5.2 Relationship to Existing Literature

Our findings complement and extend the emerging LMIC-focused LLM literature. The Pakistan trial (Qazi et al. 2026) demonstrated that AI-literate physicians can use LLMs to improve diagnostic reasoning, while the Rwanda study (Ntirenganya et al. 2026) showed that general-purpose LLMs can outperform local clinicians on quality metrics. Our study adds a new dimension: when physicians are asked to compare two AI systems directly, context-specific design produces meaningfully better assessments. This is consistent with broader concerns raised in the digital health literature about the risks of deploying AI tools developed in one context into another without adaptation (Ciecierski-Holmes et al. 2022; Wahl et al. 2018).

The finding also speaks to the know–do gap literature. If AI tools are to help close the gap between what clinicians know and what they do in practice, those tools must provide recommendations that are not only clinically accurate but also feasible, affordable, and aligned with the resources actually available in the practice setting (Das and Hammer 2014; Kruk et al. 2018). A recommendation to prescribe a drug that is not available in the local formulary, or to order a diagnostic test that the facility does not offer, is clinically irrelevant regardless of its technical accuracy. VITA’s design explicitly addresses these constraints, and our results suggest that physicians recognize and value this contextual appropriateness.

### 5.3 Implications for Policy and Practice

These findings have several implications for health systems and policymakers in LMICs. First, they suggest that informal physician adoption of general-purpose AI chatbots— while understandable given the scarcity of clinical support—should not be assumed to provide adequate support, particularly for treatment, dosing, and evidence-based prescribing decisions. Second, they underscore the value of investing in context-specific AI tools that are trained on local data and aligned with national guidelines. Third, they highlight the importance of physician-centered evaluation as a complement to purely technical benchmark assessments. The fact that physicians themselves report meaningful quality differences between the two tools speaks directly to adoption, trust, and long-term usability.

### 5.4 Limitations

Several limitations should be noted. First, the study relies on physicians’ assessments rather than patient outcomes; our findings provide insight into physicians’ perceptions of outputs from the two models, which might differ from performance in clinical settings where workflow integration challenges become salient. Future research should examine whether VITA’s perceived advantages translate into improved clinical decisions and health outcomes in routine practice. Second, the sample of 37 physicians, while spanning multiple sites and specialties, is relatively small and concentrated in India and Bangladesh. The study was not designed to be globally representative, but instead to provide a rapid assessment of providers’ assessments. Third, the evaluation was conducted via a structured survey of provider assessments among providers identified through a convenience sampling; the willingness of such participants to adopt and test AI tools might be more representative of early adopters of technology who might be AI-friendly. However, it is unlikely that AI-friendly providers would necessarily be biased towards VITA. Fourth, the ChatGPT version used (5.2) reflects a specific point in time; the performance of general-purpose LLMs continues to evolve rapidly. Finally, the study design, in which physicians evaluated both tools, may introduce carryover or comparison effects, though the regression analysis suggests these do not drive the observed differences.

### 5.5 Future Directions

As VITA is adopted by more physicians and healthcare organizations across India and similar LMIC settings, future studies will expand this evaluation in several directions. These include real-time clinical evaluations embedded in routine care workflows, assessment of patient health outcomes associated with AI-assisted decision-making, comparison of VITA’s performance against other leading AI tools used by healthcare providers (beyond ChatGPT), evaluation across additional specialties and geographic contexts, and studies of long-term adoption patterns, trust development, and integration with existing health information systems. The ultimate goal is to empirically test whether purpose-built AI-powered clinical reasoning tools lead to measurable improvements in the quality, safety, and equity of healthcare delivery in resource-limited settings.

## 6. Conclusion

In this first head-to-head physician evaluation of a purpose-built versus general-purpose AI clinical decision support tool in an LMIC setting, physicians consistently rated VITA as superior to ChatGPT across all dimensions of clinical quality. The advantage was most pronounced in evidence quality—the dimension most critical for safe prescribing in resource-limited settings—and was robust to controls for physician characteristics, case type, and evaluation order. These results provide strong evidence that contextual relevance, including local guidelines, formulary constraints, and antimicrobial resistance data, is not a secondary feature but a core determinant of AI utility in LMIC clinical settings. As healthcare systems in low-resource settings grapple with the rapid and often informal adoption of AI by clinicians, this study makes the case for investing in purpose-built tools that are designed from the ground up to meet the specific needs of these contexts.

## Data Availability

All data produced in the present study are available upon reasonable request to the authors

## Author Contributions, Acknowledgements and Conflict of Interest

CM: Conception, Development, Analysis, Drafting and Manuscript Finalization; All other co-authors: development and design of study, reviewing results, and finalization of manuscript.

VALID consortium includes physicians who volunteered to participate in evaluation of AI tools in LMIC and Indian settings. The consortium includes Charuta Mandke, Hemendra Kumar Agrawal, Bhartendu Bharti, Mayank Chansoria, Gourav Gupta, Sumit Kumar Rawat, Nirmal Kanti Sarkar, Anil Singh, Shweta Walia, Maisha Samiha Binte Akter, Syeda Tohara Tuz Alam, Anjali Bharill, Mridima Chandra, Madhu Chaudhary, Prabhakar Gupta, Shobhit Gupta, Shiva Gupta, Juhi Gupta, Parveen Hussain, Revanth K., Aman Kushwaha, Yusuf Malik, Abu Hena Mohammad Masrur, Mehedi Monwar Hossain Meham, Ankit Mishra, Suji P.S., Swaha Panda, Prashant Parmar, Rajendra Prasad Patel, Pias Paul, Aaditya A. Prabhudesai, Vinothkannan R., Nikhat Rehman, Mukesh Saket, Nihil Singh, Rishubh Pratap Singh, Murari Sravani, and Rafia Tanveer.

Charuta Mandke is co-founder of VITA. CM did not have access to primary data collected in the data and all analysis was conducted using de-identified analytical datasets.

Thanks to team members at iKites (development team for VITA), as well as VITA co-founders for support. VITA has been developed with initial funding from Social Alpha (Bangalore, India).

## References

Agrawal A, Kalyanpur A, et al. (2024). Artificial intelligence-powered healthcare for India: Promises, opportunities and challenges. National Medical Journal of India.

Alami H, Lehoux P, et al. (2020). Artificial intelligence in health care: Laying the foundation for responsible, sustainable, and inclusive innovation in low- and middle-income countries. Globalization and Health, 16(1), 52.

Ashkenazi S, et al. 2025. “Large Language Models in Real-World Clinical Workflows: A Systematic Review.” Frontiers in Digital Health 7:1659134

Ciecierski-Holmes T, et al. (2022). Artificial intelligence for strengthening healthcare systems in low- and middle-income countries: A systematic scoping review. npj Digital Medicine, 5(1), 162.

Das J, Hammer J. (2014). Quality of primary care in low-income countries: Facts and economics. Annual Review of Economics, 6, 525–553.

Das J, Holla A, Das V, Mohanan M, Tabak D, Chan B. (2012). In urban and rural India, a standardized patient study showed low levels of provider training and huge quality gaps. Health Affairs, 31(12), 2774–2784.

Eid R, et al. (2025). Global ChatGPT interest across healthcare and education access. [Journal reference].

Goh E, et al. (2024). Large language model influence on diagnostic reasoning: A randomized clinical trial. JAMA Network Open, 7(10), e2440969.

Goh E, et al. (2025). GPT-4 assistance for improvement of physician performance on patient care tasks: A randomized controlled trial. Nature Medicine, 31, 1233–1238.

Khosla V. (2023). A free primary care physician for every Indian available 24×7. The Times of India, December 31.

Kruk ME, Gage AD, Arsenault C, et al. (2018). High-quality health systems in the Sustainable Development Goals era: Time for a revolution. The Lancet Global Health, 6(11), e1196–e1252.

Leslie HH, et al. (2017). Training and supervision did not meaningfully improve quality of care for pregnant women or sick children in sub-Saharan Africa. Health Affairs, 36(9), 1555–1563.

Mohanan M, Vera-Hernández M, Das V, et al. (2015). The know-do gap in quality of health care for childhood diarrhea and pneumonia in rural India. JAMA Pediatrics, 169(4), 349–357.

National Academies of Sciences, Engineering, and Medicine. (2018). Crossing the Global Quality Chasm: Improving Health Care Worldwide. Washington, DC: The National Academies Press.

Ntirenganya F, et al. (2026). LLM-assisted clinical reasoning for community health workers in Rwanda. Nature Health.

Qazi SA, et al. (2026). Large language model diagnostic assistance for physicians in a lower-middle-income country: A randomized controlled trial. Nature Health.

Si Y, Yang Y, Wang X, et al. (2024). Quality and accountability of ChatGPT in health care in low- and middle-income countries: Simulated patient study. Journal of Medical Internet Research, 26, e56121.

Tallarico V, et al. 2025. “A Cluster Randomized Trial Assessing the Effect of a Digital Health Clinical Decision Support Algorithm on Quality of Care for Sick Children in Tanzania.” PLOS Digital Health 4 (or current volume): e0000694.

Tamrat T, et al. 2020. “Electronic Clinical Decision Support Algorithms Incorporating Point-of-Care Diagnostic Tests in Low-Resource Settings: A Target Product Profile.” BMJ Global Health 5 (2): e002067.

Wahl B, et al. (2018). Artificial intelligence (AI) and global health: How can AI contribute to health in resource-poor settings? BMJ Global Health, 3(4), e000798.

World Health Organization. (2019). WHO Guideline: Recommendations on Digital Interventions for Health System Strengthening. Geneva: WHO.

World Health Organization. (2020). Digital Health for the End TB Strategy: An Agenda for Action. Geneva: WHO.

